# Cancer diagnosis in primary care after second pandemic year in Catalonia: a time-series analysis of primary care electronic health records covering about five million people

**DOI:** 10.1101/2022.03.27.22272930

**Authors:** Núria Mora, Carolina Guiriguet, Roser Cantenys, Leonardo Méndez-Boo, Mercè Marzo-Castillejo, Mència Benítez, Francesc Fina, Mireia Fàbregas, Eduardo Hermosilla, Albert Mercadé, Manuel Medina, Ermengol Coma

## Abstract

**Background:** During COVID-19 pandemic, incidence of chronic disease had drastically been reduced due to health care interruptions. The aim of this study is to analyze cancer diagnosis during the two years of the COVID-19 pandemic.

**Methods:** Time-series study of malignant neoplasms, using data from the primary care electronic health records from January 2014 to December 2021. We obtained the expected monthly incidence using a temporary regression adjusted by trend and seasonality. We additionally compared cancer incidence in 2019 with those of 2020 and 2021 using the T-Test. We performed analysis globally, by sex and by type of cancer.

**Results:** During 2020, the incidence of cancer had reduced by -21% compared to 2019 (p-value <0.05). Greater reductions were observed during lockdown in early 2020 (>40%) and with some types of cancers, especially prostate and skin cancers (−29.6% and -26.9% respectively, p-value<0.05). Lung cancers presented statistically non-significant reductions in both years. Cancer diagnosis returned to expected around March 2021, and incidence in 2021 was similar to that of 2019 (overall difference of 0.21%, p=0.967). However, -11% reduction still was found when comparing pandemic months of 2020-2021 with pre-pandemic months (2019-2020)

**Conclusions:** Although primary care cancer diagnostic capacity in 2021 has returned to pre-pandemic levels, missing diagnoses during the last two years have not been fully recovered.

**Key messages:**

- Cancer diagnoses have dramatically dropped during 2020 worldwide.
- We observe a -21% decline in 2020, but a return to pre-pandemic diagnosis capacity in 2021.
- A -11% outstanding drop was still found comparing pre-pandemic to pandemic months.
- Reductions were greater during the lockdown (>40%).
- Lung and breast cancers presented fewer reductions while prostate and skin cancers had greater drops.
- Missing diagnoses during the last two years have not been fully recovered

## Background

The coronavirus disease 2019 (COVID-19) pandemic has put healthcare systems around the world under unprecedented pressure, leading to a disruption in health care which included suspension of screening programs, reduction of chronic disease control and reduction of non-COVID diagnosis, among others [1, 2].

Cancer diagnoses decreased during 2020 in several countries [3-6]. In a previous study in Catalonia (Spain), we observed a 34% reduction on cancer diagnosis from March to September 2020, but this reduction was even higher during the lockdown (>40%) [7]. These disruptions were likely to lead to significant delays in cancer diagnosis and treatment, which would translate into more cancers diagnosed in later stages and decreased long-term survival [4,8-10].

Although this situation is well-described in several articles, these studies are focused on cancer trends in 2020, and little is known about cancer diagnosis evolution during the second year of the COVID-19 pandemic. After the first wave of COVID-19, some restrictions were lifted in several countries and screening programmes were recovered, thus contributing to a change in cancer’s diagnostic trends. For instance, in a study in Canada, researchers observed that after a huge drop of cancer screening during the first months of COVID-19 pandemic, within a few months, programs and cancer management have been adapted to the new situation and one year later, the number of procedures reached pre-pandemic levels, suggesting that cancer diagnostic capacity could be returned to normal [11].

The aim of our study is to analyze cancer diagnosis through the analysis of primary care electronic health records (EHR) data in Catalonia almost two years after the beginning of the COVID-19 pandemic, by updating previous figures [7] and with the hypothesis that after a reduction in cancer diagnosis in 2020, during 2021 cancer incidence has reached pre-pandemic levels.

## Methods

We performed a time-series study of malignant neoplasm case reported diagnoses. Data were extracted from the primary care EHR of the Institut Català de la Salut (Catalan Institute of Health; or ICS, its Catalan initials). ICS is the main primary care provider in Catalonia. It manages around 75% of all primary care practices (PCPs) in the Catalan public health system and covers about 5.8 million people. We included all patients aged over 14 years with a clinical diagnosis of cancer according to the International Classification of Diseases 10th revision, clinical modification ICD-10CM (**Supplementary material S1**). The study period was from January 2014 to December 2021. We divided this period into two sets for the time-series analysis: training (2014-2019) and analysis (2020-2021). Validation and details of the model used in this work were described elsewhere [7].

Monthly cancer incidence rates per 100,000 inhabitants were calculated. Time-series were performed globally, and by sex and type of neoplasm. In addition, some related cancer diagnostic procedures such as mammograms and colonoscopies were also analyzed.

### Statistical analysis

We obtained the point forecast and 95% confidence interval (95%CI) of expected monthly cancer incidence rate for the analysis period by projecting the time series regression model from the incidence rate per 10^5^ inhabitants fitted in the training period. The adjustment variables were the trend and seasonality of the time series. We also calculated the percentage of reduction as follows: (observed incidence rate−expected incidence rate)/expected incidence rate. More details are provided in a previous publication [7].

Additionally, we compared the monthly average of new cancer diagnoses in 2019 with 2020 and 2021 using the independent samples T-Test and we calculated the percentage of reduction compared to 2019.

Finally, we calculated the Pearson correlation coefficient between monthly colorectal cancer (CRC) and colonoscopies and between mammograms and breast cancer during 2020 and 2021 months.

All analyses were conducted using R V.3.5.1.

## Results

From January 2014 to December 2021, 327,452 new malignant neoplasms were registered in the Catalan primary care EHR. This represents a monthly average incidence per 10^5^ inhabitants of 72.5 during the 2014-2018 period, 73.0 during 2019, only 57.4 during 2020, and 73.2 in 2021.

Figure 1. shows the observed and estimated rates of monthly new cancer diagnoses (with 95%CI) since January 2020. We observed great reductions during 2020, especially during March (−35%), April (−59%) and May (−45%), coinciding with the first wave of COVID-19 and the lockdown in Spain. After that period, incidence was below the expected but with lesser reductions (i.e - statistically significant reductions between 10-20%) until March 2021 where rates remained closer to the estimated; and slightly above the expected for women (see **Supplementary material S2**).

**Figure 1.**
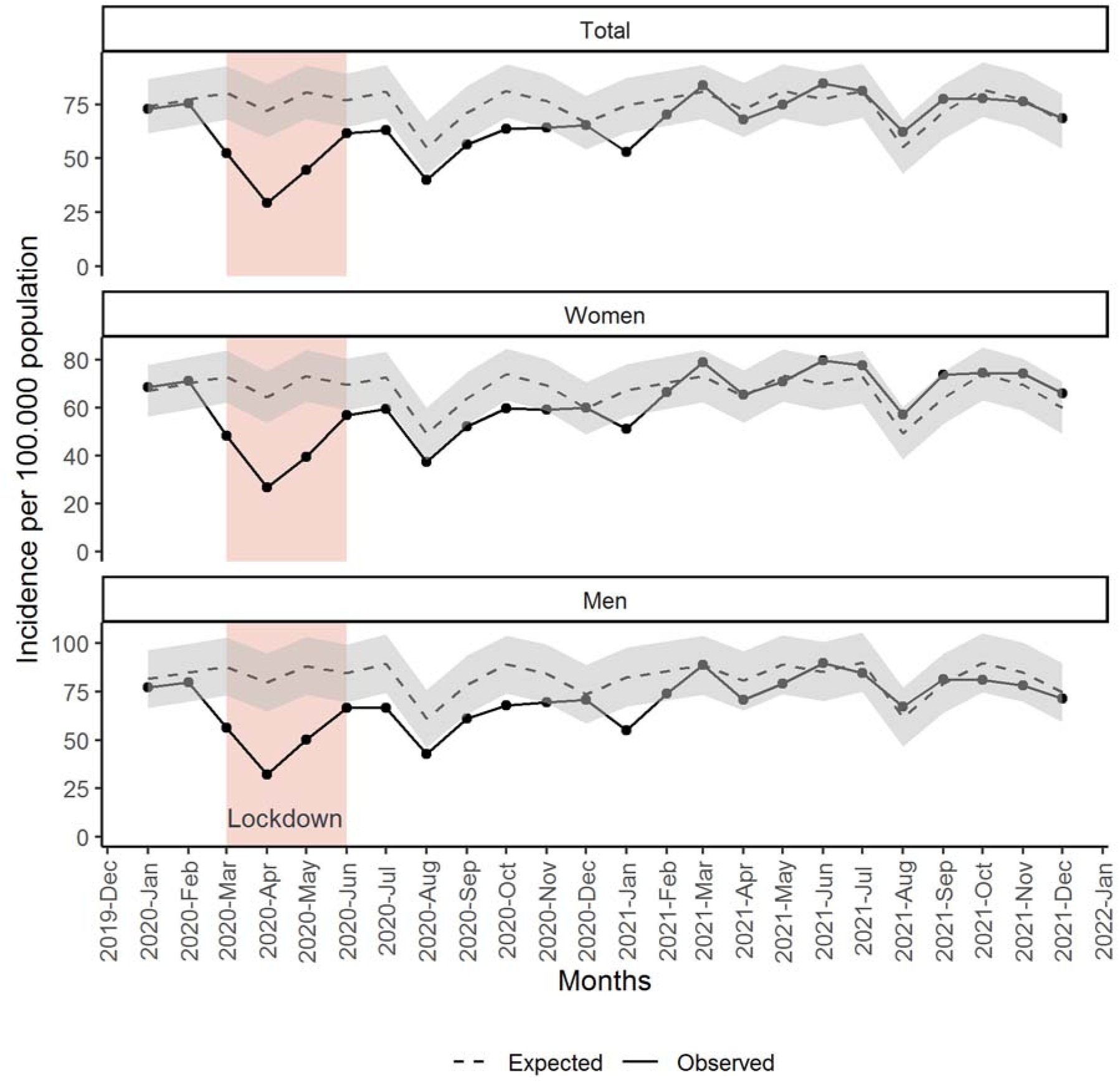
Monthly observed and expected (with 95% CI) incidence of malignant neoplasm diagnosed in Catalan primary care since January 2020, total and by sex.

During the pre-pandemic year 2019, 42,799 new malignant neoplasms were registered in the catalan primary care EHR. This number decreased to 34,024 in 2020 and slightly increased in 2021 (43,719 new cancers registered). By comparing the monthly average of new cancer diagnoses in 2019, 2020 and 2021, we observed a -21.4% reduction in 2020 (p-value <0.05) and a statistically non-significant increase of 0.21% in 2021 (p-value 0.967). However, if we compared the pandemic months of 2020 and 2021 (from March 2020 to December 2021) to the pre-pandemic period (from January 2019 to February 2020) an overall -11.8% decline was still found (p-value <0.05).

All types of cancer presented reductions in 2020, although they differ depending on the type of cancer (**Figure 2**). Reductions in 2020 compared to 2019 accounted for a -29.6%, -26.9%, - 25.7%, - 23.9%, -19.8% and -17.6% in prostate cancer, skin non-melanoma, melanoma, CRC, breast cancer and other cancers, respectively (p-values <0.05). Contrarily, lung cancer had a non-significant reduction of -6.6% (p-value=0.266). During 2021, all cancers reached similar incidences to those of 2019, although prostate and melanoma had -11.2% (p-value =0.12) and - 6.7% (p-value=0.34) drops, respectively; while breast cancer had a 7% increase (p-value=0.19).

**Figure 2.**
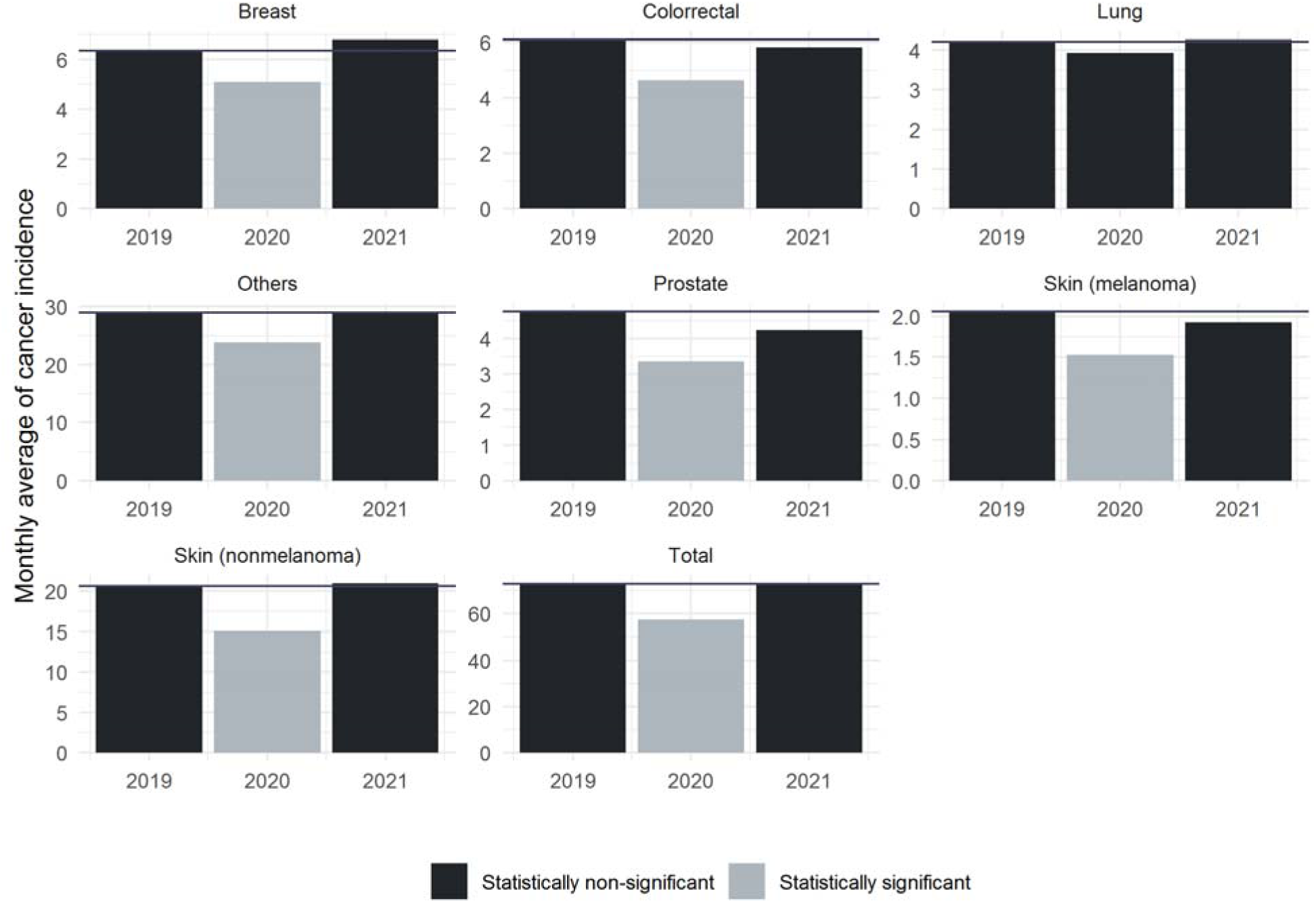
Mean monthly incidence of cancer by type of cancer and year (2019-2021). Statistically significant differences are shown in lighter color

Finally, we observed a high correlation during 2020 and 2021 between CRC and colonoscopies with a R^2^ of 0.78, and between mammograms and breast cancer R^2^ = 0.73. Number of diagnostic procedures by year are provided in **Supplementary material S3**.

## Discussion

We hereby describe a reduction of cancer diagnosis during 2020 and a return to the expected during 2021. The reduction was greater during the early stages of the pandemic, coinciding with the lockdown in Spain. However, after the first months, the incidence was also consistently under the expected until March 2021, just after the third COVID-19 wave in Spain, where diagnosis returned to baseline levels (i.e. comparable to 2019) and remained relatively stable thereafter. Other studies worldwide described similar reductions during 2020 as in our work [3-6, 12], highlighting the impact of the pandemic to non-COVID diseases. Several reasons have been described to explain this decrease, such as disruption of health care due to the pandemic, drop of face-to-face visits, halt of screening and diagnostic procedures and non-essential services, some COVID-19 control measures, and changes in patients’ health-seeking behavior, among others [7, 13-14].

Nevertheless, primary care adapted quickly to the COVID-19 pandemic within a few months, and cancer incidences in 2021 returned to similar values to those of 2019 in Catalonia. Despite that, the return to pre-pandemic diagnostic level in 2021 did not compensate for the drops in 2020. Although a part of this reduction could be related to the harvest effect due to the high impact of COVID-19 first wave in Spain in terms of mortality, a study performed in Catalonia during 2020 estimated that only a 4% of diagnosis may have been lost to COVID-19 deaths [2]. This means that there has yet to be a complete recovery with a reduction of diagnosis, still outstanding. This could still have consequences described elsewhere such as more advanced diagnosis and delays in treatments that could affect patients’ survival [9, 15-17]. This return to 2019 diagnostic capacity is not yet described in many articles, although some studies observed that screening programmes and cancer diagnosis returned close to pre-pandemic levels late 2020 [11, 12].

Limitations of this analysis include ecological analysis, the use of EHR data, and alterations in patients’ healthcare-seeking behavior during COVID-19 pandemic.

In conclusion, although primary care cancer diagnostic capacity in 2021 has returned to pre-pandemic levels, lost diagnoses during the last two years have not been fully recovered.

Additional strategies may be needed to address remaining backlogs and potential consequences in terms of long-term cancer survival.

## Additional sections

## Ethics approval

This study was done in accordance with existing statutory and ethical approvals from the Clinical Research Ethics Committee of the IDIAPJGol (project code: 20/172-PCV).

## Data availability statement

EHR data and analytical code are provided at https://github.com/ErmengolComa/cancer2021/

## Acknowledgments

We would like to acknowledge the efforts of all members of the SISAP team during the last months. We would also like to thank all the primary care healthcare professionals in Catalonia during these challenging times.

