## Supplementary material for "Cancer diagnosis in primary care after second pandemic year in Catalonia: a time-series analysis of primary care electronic health records covering about five million people"

**Supplementary material S1.** ICD-10 codes and ICPC-2 codes used to identify malignant neoplasms

| **Type of malignant neoplasm** | | **ICPC-2 Code** | **ICD 10 Code** |
| --- | --- | --- | --- |
| Colorectal | | D75 | C18-C21 |
| Lung | | R84 | C33, C34, C46.5 |
| Skin non-melanoma | | S770 | C44, C46.0 |
| Melanoma | | S771 | C43 |
| Breast (women) | | X76 | C50 |
| Prostate | | Y77 | C61 |
| Others | Stomach | D74 | C16 |
|  | Pancreas | D76 | C25 |
|  | Other digestive organs | D77 | C17, C46.2, C46.4, C00-C08, C15, C17, C22, C23, C24, C26, C45.1, C46.2, C46.4, C48 |
|  | Brain and other parts of nervous system | N74 | C47, C70, C71, C72 |
|  | Cervix uteri | X75 | C53 |
|  | Other female genital organs | X77 | C51, C52, C54-C57 |
|  | Lymphoma | B72 | C81-C86 |
|  | Leukemia | B73 | C91-C95 |
|  | Other hematopoietic and related tissue | B74 | C88, C90, C96, C26.1, C37, C46.3 |
|  | Bladder | U76 | C67 |
|  | Kidney and renal pelvis | U75 | C64, C65 |
|  | Ureter and other urinary organs | U77 | C66, C68 |
|  | Thyroid | T71 | C73 |
|  | Other respiratory organs | R85 | C09-C14, C30-C32, C39, C45.0 |
|  | Bone and articular cartilage | L71 | C40, C41, C46.1, C49 |
|  | Breast and genital organs in men | Y78 | C50, C60, C62, C63 |
|  | Ear | H75 | C30.1 |
|  | Eye | F74 | C69 |
|  | Heart | K72 | C38.0, C45.2 |
|  | Related to pregnancy | W72 | C58 |
|  | Malignancy non specified | A79 | C14, C30, C38, C39, C45, C45.7, C45.9, C46, C46.7, C46.9, C4A, C76, C77, C78, C79, C7A, C7B, C80 |

**Supplementary material S2** Monthly percentage of change (with 95% CI) of observed cancer diagnosis compared to the expect by month

| Period | Total | Women | Men |
| --- | --- | --- | --- |
| January 2020 | -1.87%  (-15.94% - 17.86%) | 2.33%  (-11.88% - 22.02%) | -5.51%  (-20.2% - 15.81%) |
| February 2020 | -2.6%  (-16.07% - 16.03%) | 1.35%  (-12.2% - 19.83%) | -6.03%  (-20.17% - 14.18%) |
| March 2020 | -34.79%  (-43.53% - -22.86%) | -33.68%  (-42.24% - -22.14%) | -35.78%  (-45.15% - -22.53%) |
| April 2020 | -59.1%  (-65.11% - -50.57%) | -58.62%  (-64.57% - -50.27%) | -59.51%  (-65.92% - -50.13%) |
| May 2020 | -44.55%  (-51.95% - -34.45%) | -46.25%  (-53.16% - -36.94%) | -43.09%  (-51.37% - -31.41%) |
| June 2020 | -19.74%  (-30.89% - -4.29%) | -18.26%  (-29.24% - -3.24%) | -21.05%  (-32.97% - -3.97%) |
| July 2020 | -22.06%  (-32.44% - -7.92%) | -18%  (-28.63% - -3.65%) | -25.54%  (-36.25% - -10.5%) |
| August 2020 | -27.05%  (-40.5% - -5.75%) | -23.59%  (-37.39% - -1.97%) | -30.01%  (-43.83% - -7.16%) |
| September 2020 | -20.31%  (-32.17% - -3.43%) | -18.31%  (-30.14% - -1.67%) | -22.04%  (-34.57% - -3.58%) |
| October 2020 | -21.65%  (-32.03% - -7.52%) | -19.3%  (-29.6% - -5.47%) | -23.72%  (-34.74% - -8.22%) |
| November 2020 | -16.12%  (-27.81% - 0.09%) | -14.5%  (-26.03% - 1.29%) | -17.55%  (-30.03% - 0.35%) |
| December 2020 | -1.91%  (-17.33% - 20.59%) | 0.28%  (-15.09% - 22.43%) | -3.81%  (-20.1% - 20.83%) |
| January 2021 | -28.9%  (-39.17% - -14.45%) | -23.82%  (-34.51% - -8.96%) | -33.26%  (-43.69% - -18.1%) |
| February 2021 | -9.71%  (-22.3% - 7.74%) | -5.41%  (-18.18% - 12.08%) | -13.45%  (-26.54% - 5.31%) |
| March 2021 | 3.63%  (-10.36% - 22.79%) | 7.81%  (-6.25% - 26.84%) | -0.03%  (-14.7% - 20.73%) |
| April 2021 | -6.14%  (-20.05% - 13.62%) | 1.06%  (-13.61% - 21.74%) | -12.22%  (-26.19% - 8.26%) |
| May 2021 | -7.58%  (-20.01% - 9.42%) | -3.57%  (-16.11% - 13.36%) | -11.09%  (-24.1% - 7.29%) |
| June 2021 | 9.34%  (-5.97% - 30.61%) | 13.89%  (-1.58% - 35.11%) | 5.37%  (-10.62% - 28.32%) |
| July 2021 | -0.24%  (-13.63% - 18.06%) | 6.55%  (-7.41% - 25.46%) | -6.04%  (-19.63% - 13.09%) |
| August 2021 | 12.09%  (-8.68% - 45.09%) | 15.94%  (-5.19% - 49.18%) | 8.76%  (-12.76% - 44.37%) |
| September 2021 | 8.27%  (-7.95% - 31.44%) | 15.02%  (-1.8% - 38.79%) | 2.49%  (-14.05% - 26.91%) |
| October 2021 | -4.79%  (-17.5% - 12.56%) | 0.68%  (-12.31% - 18.19%) | -9.56%  (-22.7% - 8.95%) |
| November 2021 | -1.25%  (-15.12% - 18.04%) | 6.81%  (-7.74% - 26.82%) | -8.21%  (-22.17% - 11.86%) |
| December 2021 | 2.27%  (-13.91% - 25.94%) | 9.79%  (-7.2% - 34.39%) | -4.13%  (-20.43% - 20.56%) |

**Supplementary material S3** Number of colonoscopies and mammograms by year

| Year | Colonoscopies | Mammograms |
| --- | --- | --- |
| 2014 | 39330 | 69271 |
| 2015 | 45025 | 66816 |
| 2016 | 47718 | 65230 |
| 2017 | 48090 | 65734 |
| 2018 | 50023 | 63452 |
| 2019 | 53396 | 63272 |
| 2020 | 38049 | 43536 |
| 2021 | 53283 | 55145 |
